# Effect of NHS surgical hubs on elective hip-and-knee replacement volume, length of stay and waiting times: national longitudinal difference-in-differences study

**DOI:** 10.64898/2026.04.21.26351383

**Authors:** Jinglin Wen, Zecharias Anteneh, Adriana Castelli, Andrew Street, Nils Gutacker, Arabella Scantlebury, Karen Glerum-Brooks, Simon Davies, Karen Bloor, Amar Rangan, Ana Castro Avila, Pete Lampard, Joy Adamson, Peter Sivey

## Abstract

**Background:** Elective surgical hubs have been introduced across England to increase surgical capacity, improve productivity and reduce waiting times. Evidence on their effects remains limited, particularly beyond the early period of implementation. We evaluated the effect of surgical hubs on surgical volume, patient waiting times and length of hospital stay for elective hip and knee replacements in the English NHS.

**Methods:** We conducted a retrospective longitudinal study using Hospital Episode Statistics for NHS acute trusts in England from April 2014 to September 2024. The analysis included 76 trusts, of which 29 opened a surgical hub providing trauma and orthopaedic surgery during the study period and 47 did not. Outcomes were measured quarterly at trust level and comprised the number of elective hip and knee replacements, mean length of stay and mean waiting time from addition to the waiting list to admission. We used a difference-in-differences approach for staggered hub opening, with trust and quarter fixed effects and standard errors clustered at trust level.

**Results:** Surgical hub opening was associated with an increase of 43.75 hip and knee replacement procedures per quarter (95% CI 22.22 to 65.28), equivalent to a 19.1% increase relative to the pre-hub mean. Mean length of stay was 0.32 days lower following hub opening (95% CI −0.48 to −0.16), equivalent to a 7.8% reduction. There was no statistically significant overall change in mean waiting time (−14.96 days, 95% CI −33.11 to 3.19).

**Conclusions:** Surgical hub opening was associated with increased volumes of hip and knee replacement surgery and shorter hospital stays. We found no statistically significant overall reduction in waiting times. The findings suggest that surgical hubs can increase elective surgical activity, although additional capacity may not translate directly into shorter waiting times.

**Trial registration:** Not applicable.

## Introduction

Waiting times for elective surgeries have long been a substantial barrier to access in publicly funded healthcare systems. In England, waiting times in the National Health Service (NHS) have increased steadily since 2010, with a sharp escalation during the COVID-19 pandemic due to the disruption in elective care services. In response, policymakers have explored new service delivery models to expand surgical capacity and enhance productivity. Reducing elective waiting times is a core ambition of the current UK government [1], which set an interim target of treating 65% of patients within 18 weeks by March 2026, progressing toward the NHS Constitution standard of 92% by March 2029.

Elective surgical hubs have been introduced across England to improve surgical throughput and reduce waiting times. Initially introduced in 2020 in response to the COVID-19 pandemic, hubs have since been implemented more widely [2, 3]. Surgical hubs are ring-fenced facilities dedicated to elective care, either in stand-alone premises, or integrated into existing hospital sites [4]. By consolidating staff and resources while shielding elective services from emergency care pressures, surgical hubs aim to minimise cancellations and delays, thus optimising theatre and ward utilisation [5]. These efficiencies aim to improve surgical workflows and increase productivity [4]. By mid-2024, 108 hubs had been established in England, with 26 more planned by the end of 2025 [6].

Surgical hubs build on earlier models of health care delivery in England such as NHS and independent sector treatment centres (TCs), first introduced in 2003. TCs were introduced to reduce waiting times [7] and by late 2007, nearly 100 treatment centres were in operation, about half privately run [8,9]. Evidence showed that treatment centres achieved lower length of stay for low-complexity patients [8, 10, 11], but there was no clear evidence of overall cost savings. While there was some evidence of patients treated in TCs having lower waiting times, it was unclear if falls in waiting times overall could specifically be attributed to TCs [12, 13, 14]. Studies found no differences in patient health outcomes between private and public providers [11] and found that private provider entry increased the overall number of procedures performed in an area [14]. Some former NHS TCs have been redesignated as surgical hubs, showing that the programme partly continues existing arrangements.

Globally, specialised elective care centres known sometimes as ambulatory surgical centres (ASCs) [15] or day surgery units [16], are increasingly common. Evidence highlights their cost-effectiveness and efficiency benefits [17, 18, 19, 20, 21], as well as their ability to expand the market for planned surgery [22]. However, there is no definitive evidence of their benefits within the UK healthcare system.

A previous study found that early examples of new surgical hubs in the English NHS led to a 21.9% increase in high-volume, low-complexity elective surgery but no change in length of stay [23]. Existing surgical hubs were found to improve the recovery in volume from the COVID-19 pandemic and have slightly shorter length of stay. The study did not include waiting times as an outcome and only included data up until December 2022. A recent rapid review examined the impact of surgical hubs [24], finding limited evidence of beneficial effects, mainly because of focusing on their early adoption during the COVID-19 pandemic. Consequently, the hub programme lacks high-quality evidence, especially from more recent years, to support its widespread implementation.

We hypothesised that surgical hubs would increase surgical volume and reduce average length of stay by ringfencing elective capacity to achieve more efficient elective care. We expected waiting times to fall if the additional capacity translated into shorter waiting lists, although this effect might be mitigated by increases in demand associated with the opening of new facilities.

This study evaluated the effects of surgical hubs within NHS trusts on surgical volume, length of stay, and waiting times, analysing data from April 2014 to September 2024, for patients undergoing elective hip or knee arthroplasty. It focuses on hip and knee replacements, given the prominence of orthopaedic surgery in early hub implementations and the standardised nature of these procedures across providers. This is the first quantitative study as part of MEASURE, the only independent national mixed-methods evaluation of the surgical hubs programme [25].

## Methods

### Data Sources

A retrospective cohort study was conducted using two data sources. The Hospital Episode Statistics (HES) Admitted Patient Care (APC) dataset provided comprehensive records of all NHS-funded patients treated in England, including trust codes, dates when patients were added to the waiting list, admission and discharge dates, ICD-10 diagnosis codes, and procedure codes. The surgical hub database, supplied by the Department of Health and Social Care, contained hub names, trust codes, opening dates, and clinical specialities.

### Inclusion Criteria

The intervention group comprised NHS trusts that opened a surgical hub treating trauma and orthopaedic (T&O) patients between 01/03/2020 and 30/09/2024. The control group comprised T&O departments in trusts that had not yet opened a surgical hub over this period. Opening a surgical hub did not mean that all elective activity in a trust was transferred to that facility, as some elective procedures could still be undertaken elsewhere in the trust. Trusts in the control group did not open a dedicated elective-only orthopaedic facility labelled as a surgical hub during the study period, though they may have operated other elective-focused facilities not classified as hubs.

Trusts were excluded from the analysis if they had facilities similar to surgical hubs prior to March 2020, such as pre-existing NHS-owned treatment centres that were subsequently re-designated as hubs. For NHS trusts that introduced multiple surgical hubs during the study period, classification to the intervention group was based on the earliest hub that conducted T&O surgeries and opened between March 2020 and September 2024. A trust-quarter was considered to have an open hub if the hub was operational at any time during that quarter, even if it was open for only part of the quarter.

The HES APC data contain pseudonymised patient records for all publicly funded care provided in England. We analysed data for all publicly funded patients undergoing elective hip and knee replacements treated in NHS trusts from April 2014 to September 2024. Procedures were identified using OPCS-4 procedure and anatomical site codes. Hip procedures included total prosthetic replacement of the hip joint (W37–W39), prosthetic replacement of the head of femur (W46–W48), hybrid prosthetic replacement of the hip joint (W93–W95), other hip replacements (W52–W54 combined with site codes Z75.6, Z76.1 or Z84.3), and hip resurfacing arthroplasty (W58.1 and W58.2). Knee procedures included total prosthetic replacement of the knee joint (W40–W42), unicondylar or unicompartmental knee operations (W52–W54 combined with site codes Z76.5, Z77.1, Z77.4, Z84.4 or Z84.5), and hybrid prosthetic replacement of the knee joint (O18).

We excluded trusts that performed fewer than 1,000 procedures over 10 years^1^ or failed to report data for at least 41 of the 42 quarters. This ensured we focused on providers regularly performing high volumes of surgery and removed those with inconsistent reporting that could distort the results.

The population of NHS trusts comprised 124 trusts without existing surgical hubs completing at least one hip/knee replacement. This reduced to 109 trusts who reported more than 1,000 procedures over a 10 year period, and then to 76 trusts that also reported data for at least 41 of 42 quarters in the sample period. We assessed sensitivity to this sample restriction by also estimating the models without this restriction imposed.

### Outcomes

Three outcomes were examined: surgical volume, average waiting times, and average length of stay (LOS). A trust-quarter dataset was constructed by calculating the number of elective hip and knee replacements carried out in each trust per quarter. For each trust-quarter, the average length of stay (days between hospital admission and discharge) and the average waiting time (days between being added to the waiting list and admission for surgery) were also calculated.

Patients were excluded as outliers if their length of stay or waiting time exceeded the third quartile by more than three times the interquartile range (IQR). Outliers were excluded because extremely long lengths of stay or waiting times are often due to coding errors or unusual care pathways.

### Statistical analysis

The impact of the surgical hubs programme was estimated using a difference-in-differences approach, utilising the staggered introduction of hubs across NHS trusts between March 2020 and September 2024. The unit of analysis was the trust-quarter.

The primary analysis compared the change in outcomes after hub introduction in the intervention group trusts with the outcomes that would have been expected in those trusts had a hub not opened. This analysis controlled for trust-level fixed effects and quarter fixed effects, accounting for both unobserved time-invariant differences between trusts and common temporal patterns affecting all trusts in equal ways. To account for the possibility of differences in treatment effect for early and late adopters of surgical hubs, and changes in the treatment effect after the initial introduction of the hub, we implemented an estimation approach using the recently developed estimator by Borusyak, Jaravel and Spiess (BJS) (26). This method provided estimates of the average treatment effect of surgical hub opening for trusts that opened a hub for up to eight quarters after opening, presented in an event-study plot with eight quarters before and after opening. Intervention trusts contributed only observations within this event window, while all available observations from control trusts were retained to estimate untreated outcomes. Trusts opening hubs near the end of the study period contributed fewer than eight post-opening quarters because follow-up ended in September 2024. Throughout, standard errors were clustered at the trust level to account for correlation in errors within trusts.

Further methodological details are provided in Appendix A. Sensitivity analyses assessed alternative model specifications using two-way fixed effects (TWFE), removal of the main sample restrictions, adjustment for trust-level patient casemix, and exclusion of the peak COVID-19 period (2020/21) with results provided in Appendix B. All analyses were conducted using Stata 18.

### Patient and Public Involvement

Patients and carers were first involved during the design of the wider MEASURE project, providing feedback on the broad aims and methods. During this study, through workshops and follow-up discussions, they helped prioritise the focus on outcomes that reflect access and experience for people awaiting joint replacement, particularly waiting time and length of stay, and supported the decision to report these alongside procedure volume. They also enhanced our interpretation of the outcomes. For example, while lower length-of-stay may be cost-saving, patients indicated that being forced to go home very soon after a major surgery is not always desirable.

## Results

### Descriptive statistics

In the analytical period (April 2014–September 2024), 29 NHS trusts opened a surgical hub (intervention group) in the period between March 2020 and September 2024 (Figure 1) and 47 trusts did not (controls). Overall, trusts that opened hubs performed on average 228 elective hip/knee procedures per quarter compared with 154 procedures per quarter in control trusts (Table 1). Mean length of stay across intervention group trust-quarters was 3.8 days versus 3.9 days in controls. Average waiting time was 165 days in intervention group trusts and 170.0 days in controls.

**Figure 1.**
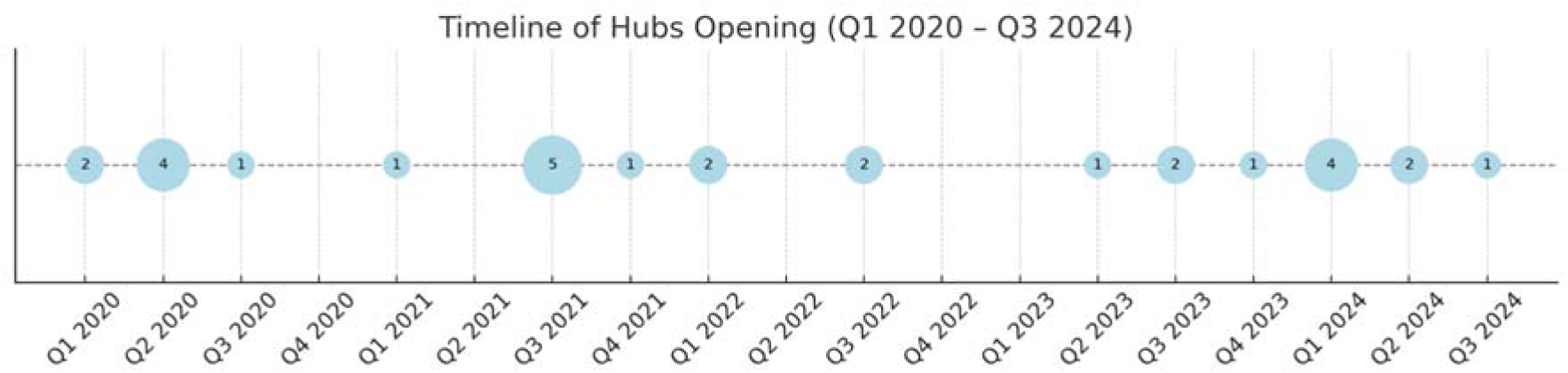

**Table 1:**
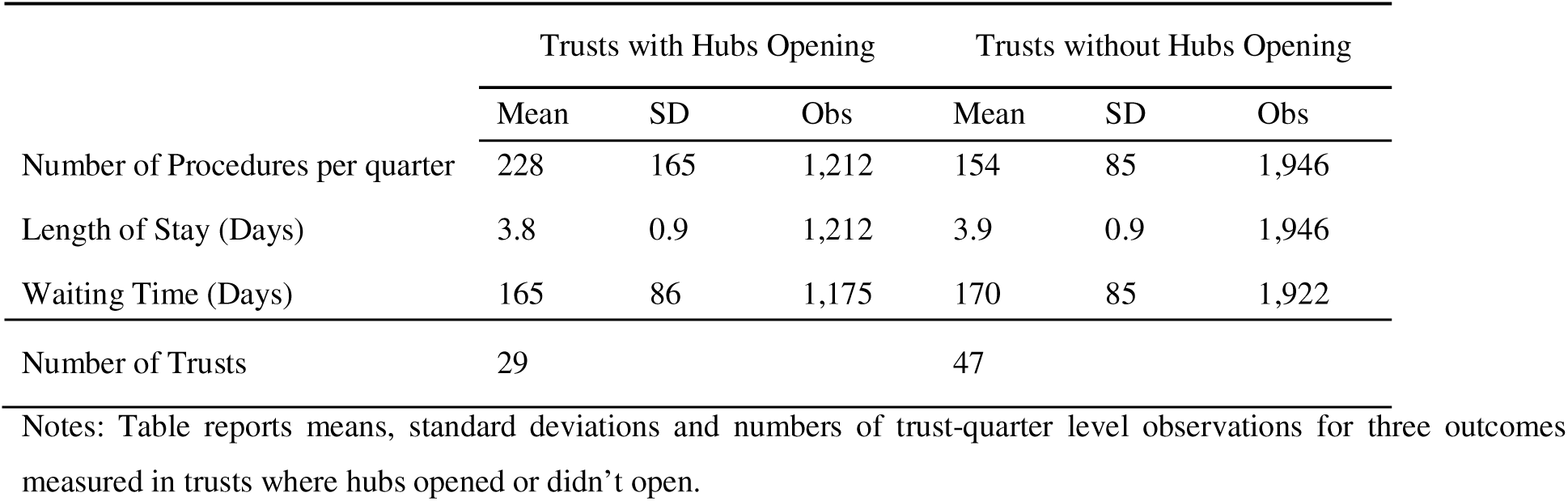
Summary Statistics.

### Estimated impacts of surgical hubs

The introduction of a surgical hub was associated with an additional 44 procedures performed per quarter (average treatment effect on the treated ATT = +43.75, 95% CI [22.22, 65.28]) and a reduction in mean length of stay of 0.32 days (ATT = −0.32, 95% CI [−0.48, −0.16]) (Table 2). These changes represent an increase of 19.1% in surgeries and a reduction of 7.8% in length of stay, compared to pre-period trust-level means. No statistically significant change in waiting time was observed following hub implementation (−14.96 days, 95% CI: −33.11 to 3.19).

**Table 2:** Impacts of surgical hub opening on trust-level outcomes.

|  | Procedures | LOS | Waiting Time |
| --- | --- | --- | --- |
| Treatment Effect | 43.75*** | -0.32*** | -14.96 |
|  | [22.22, 65.28] | [-0.48, -0.16] | [-33.11, 3.19] |
| Outcome Mean | 162 | 3.8 | 178 |
| Observations | 2,355 | 2,355 | 2,312 |

Notes: Estimates of Average Treatment Effect on the Treated (ATT) using the BJS model. Model only uses observations from intervention trusts 8 quarters before and after the intervention. This comprised 1,946 control and 409 intervention observations for procedure volume and length of stay, giving 2,355 observations, and 1,922 control and 390 intervention observations for waiting time, giving 2,312 observations. explaining the smaller sample size compared to Table 1. 95% confidence intervals in brackets. \*p < 0.1; \*\*p < 0.05; \*\*\*p < 0.01.

### Event study plots

The event study plots show estimates of the Average Treatment Effect on the Treated (ATT): the average association of the intervention with the outcomes in each quarter pre and post-intervention. The event study plot showed no evidence of differential volume trends between intervention and control groups before the hub opening, but an average increase in the intervention group of 30 to 60 procedures per quarter after hub opening (Figure 2).

**Figure 2.**
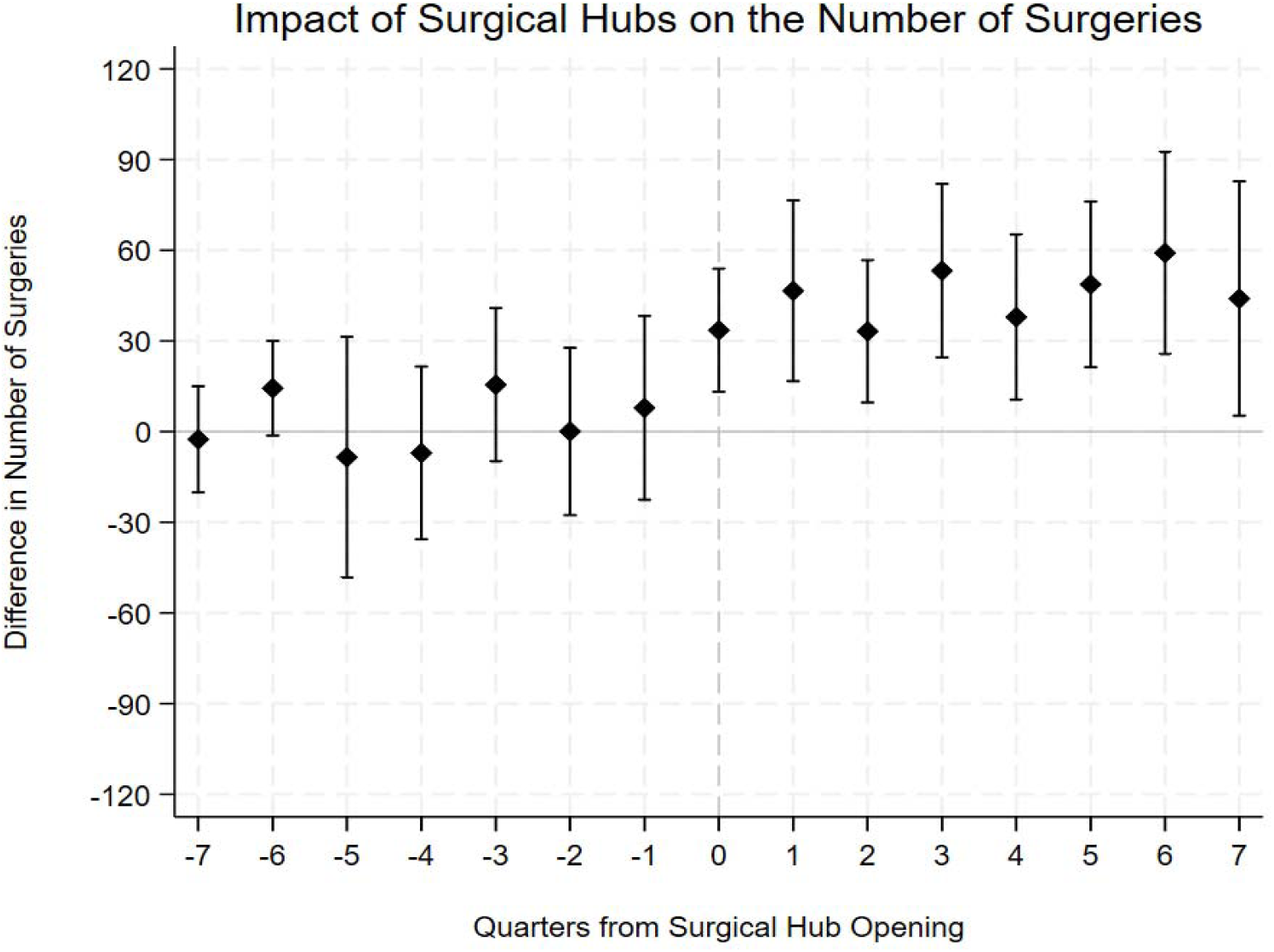
Notes: Figure presents the event-study plot of the effect of surgical hub opening on quarterly volume of procedures at the trust level using the BJS (2024) method. Bands indicate 95% confidence intervals.

The event study plot showed no statistically significant evidence of differential trends between intervention and control groups in LOS prior to hub opening, but there was some suggestive evidence of a pre-existing downward trend, which would invalidate the assumptions of the difference-in-differences approach for this outcome. Following hub opening, the analysis showed a statistically significant reduction in LOS, with the association becoming more pronounced over successive quarters (Figure 3).

**Figure 3.**
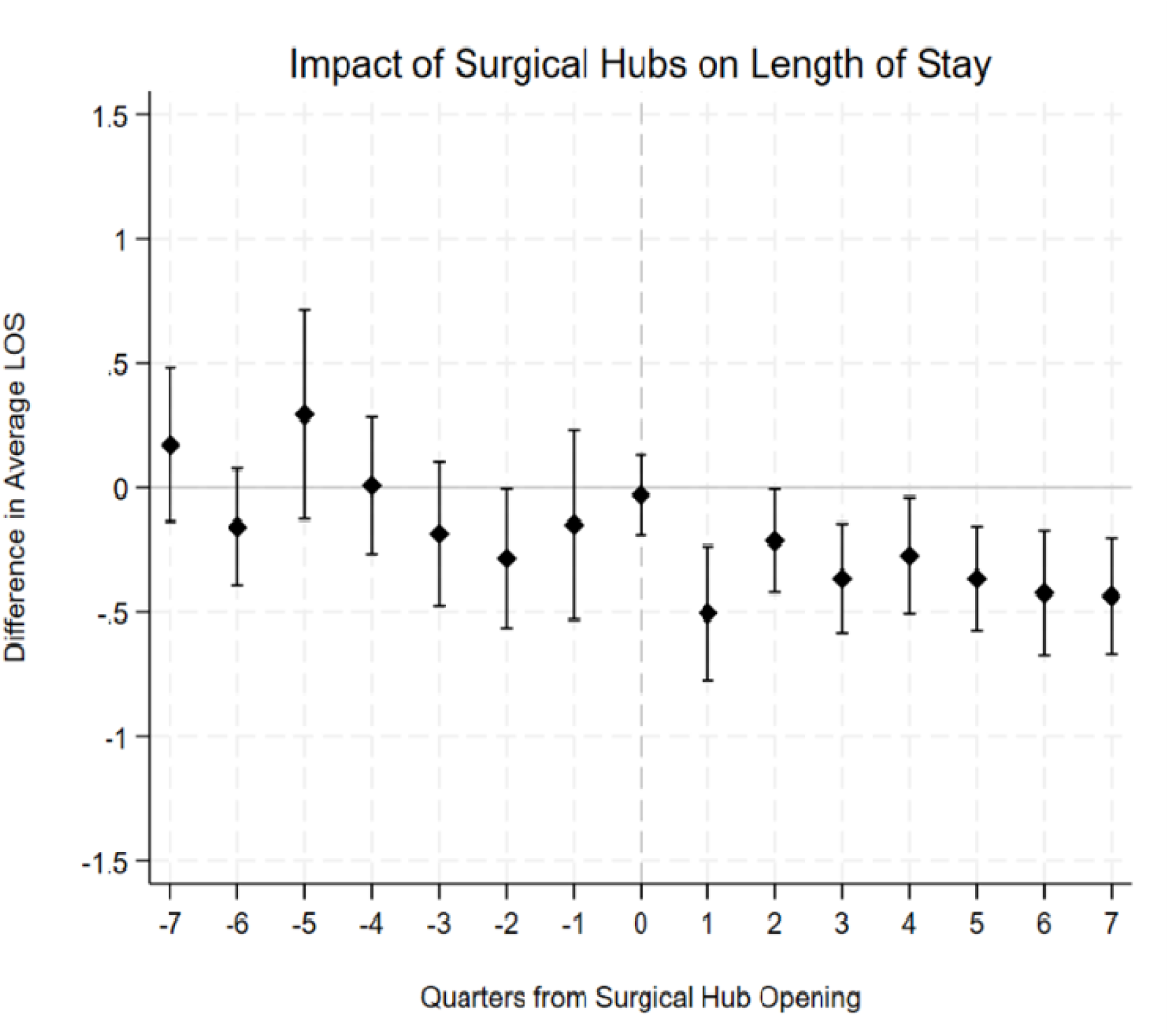
Notes: Figure presents the event-study plot of the effect of surgical hub opening on quarterly mean length of stay at the trust level using the BJS (2024) method. Bands indicate 95% confidence intervals.

The event study plot for waiting times (Figure 4) showed some evidence of differential pre-trends between intervention and control groups before hub opening with treated trusts appearing to have longer waiting times in the pre-period. There is no change in waiting times for the first 4 quarters after hub opening. While waiting times appeared to fall in quarters 4-7 after hub opening compared to control trusts, this pattern may be consistent with a return towards pre-existing trends rather than a treatment effect.

**Figure 4.**
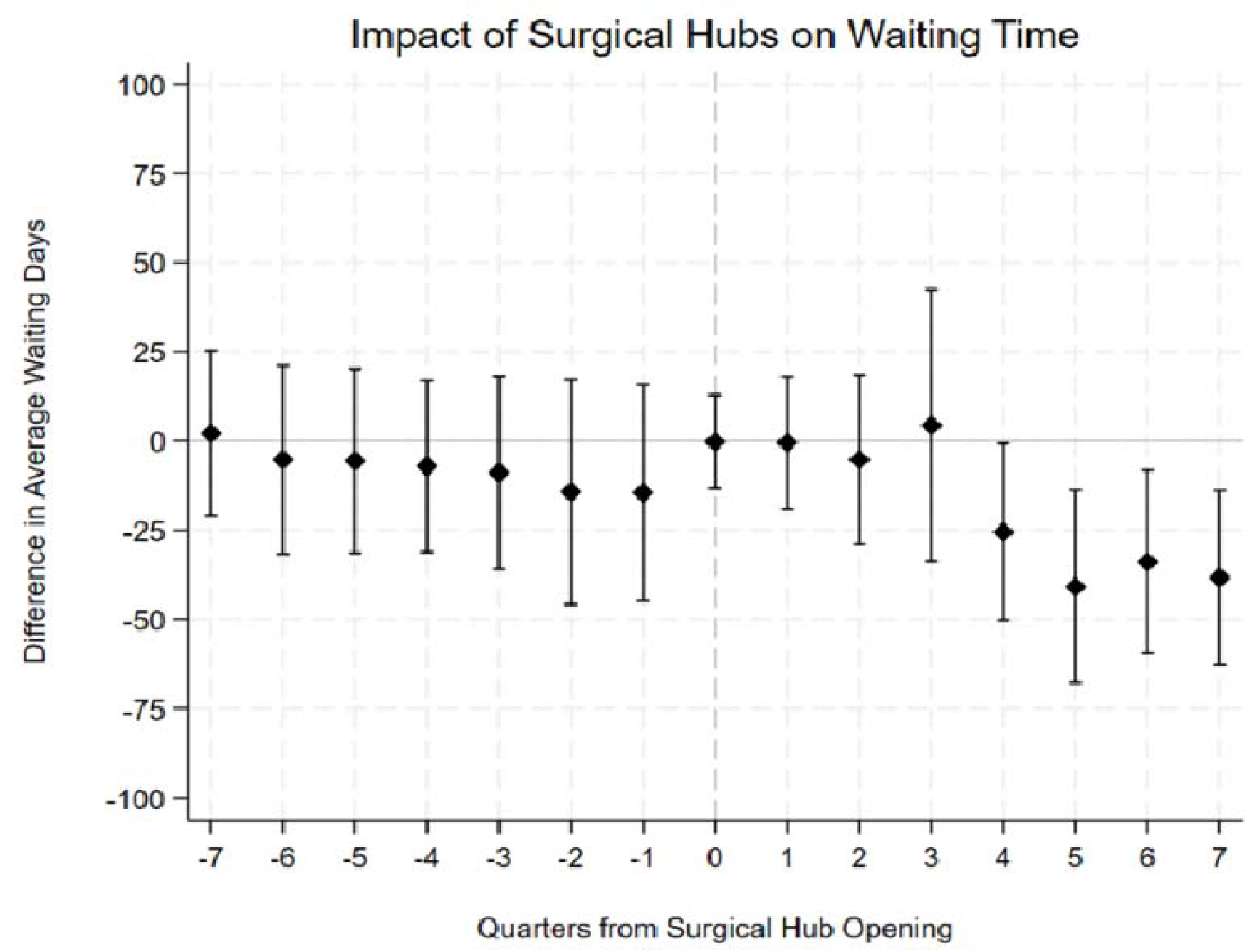
Notes: Figure presents the event-study plot of the effect of surgical hub opening on quarterly mean inpatient waiting time at the trust level using the BJS (2024) method. Bands indicate 95% confidence intervals.

Sensitivity analysis of these results is presented in Appendix B, including alternative model specifications (Table B1 and B2), analyses that include trusts with previously excluded unreliable data (Table B3 and B4), and analyses that controlled for average trust-level casemix, including age, gender, deprivation, prior emergency admissions, Charlson comorbidity, and share of procedures that were knee replacements (Table B5). We also performed a sensitivity analysis excluding data from the peak of the COVID-19 pandemic, between April 2020 and March 2021 and the estimated associations with outcomes were broadly consistent with our primary findings (Table B6). The results of the alternative specifications are summarised in Figure 5.

**Figure 5.**
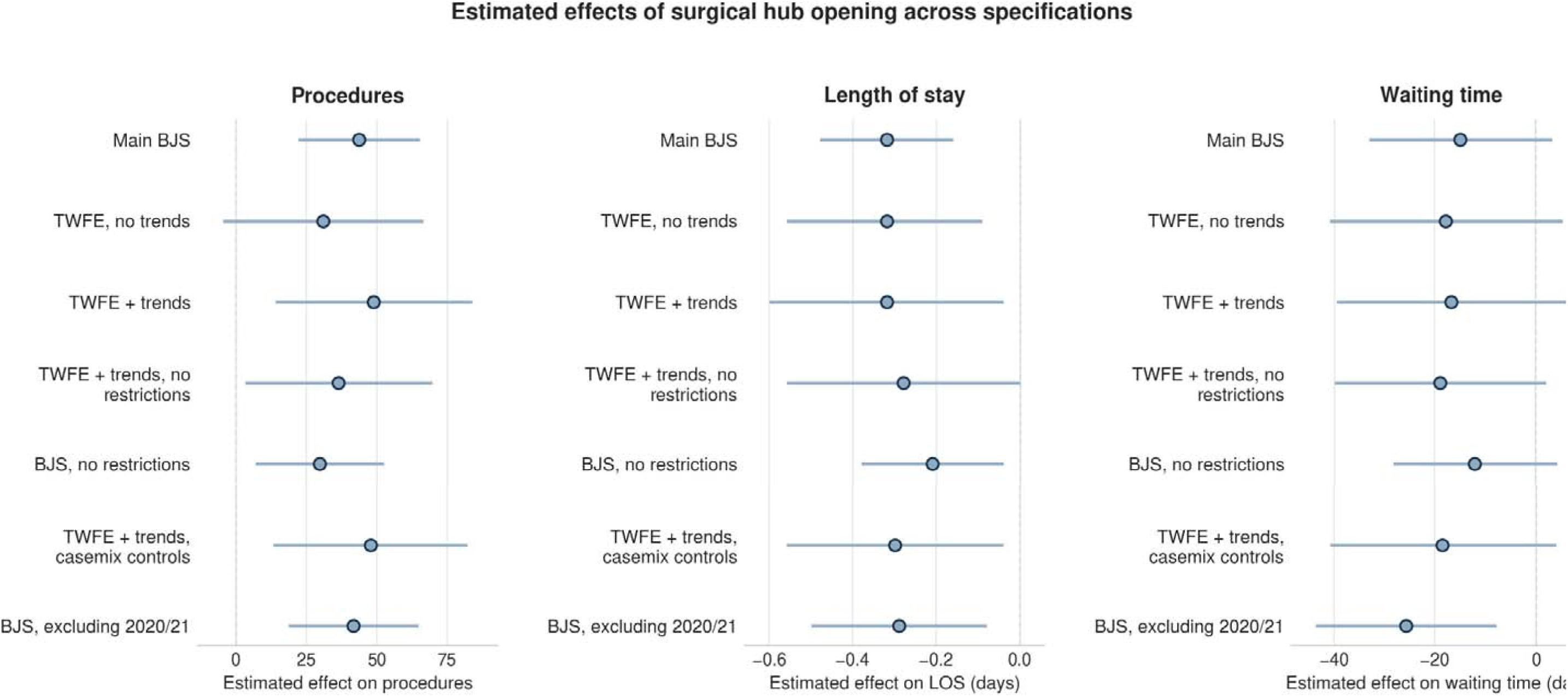
Notes: The figure shows estimated effects of surgical hub opening on trust-level outcomes in the main BJS model (Table 2) and the alternative specifications reported in Tables B1–B6. C show point estimates; horizontal lines show 95% confidence intervals.

## Discussion

This is the first study that has evaluated the association of surgical hubs opening with volume of surgeries, LOS and waiting time in the English NHS. The results showed that the introduction of surgical hubs was associated with increased surgical volume (+19.1%), reduced length of stay (−7.8%) but had no significant association with waiting times for hip and knee replacement operations in intervention trusts. We observed an immediate step-change in surgical volume that was sustained over time, whereas the estimated effect on length of stay emerged more gradually over time following the opening of the surgical hub.

The absolute increase of 44.75 hip and knee replacement procedures per quarter could be considered small when compared to a single operating theatre’s total capacity of 15 to 25 surgeries per week. However, this study measured only hip and knee replacements, which represent a fraction of the total activity a surgical hub is likely to undertake. Furthermore, the measured increase represents an increase of close to 4 joint replacements per week, which is close to a day’s standard work for joint arthroplasty, tying in well with the relative increase of 19.1% for these specific procedures.

The estimated association of hub opening with LOS was relatively small and there was some visual evidence of a pre-existing trend of falling LOS in trusts that opened hubs compared to those that did not, therefore we must treat this estimate with caution. For waiting times, the results did not show a statistically significant overall fall in hub-opening trusts. The event study showed an apparent fall in waiting times in the second year after hub implementation, but this pattern may be consistent with a return to pre-existing trends rather than a causal effect of hub opening.

### Comparison with other studies

Few previous studies have evaluated the effect of surgical hubs or similar facilities on hospital performance measures. The other comparable study of surgical hubs, which used a synthetic control method and included all surgical specialties, found a 21.9% increase in surgical activity following the introduction of surgical hubs [23], broadly comparable to our main findings. However, their analysis only used data up to December 2022 and did not assess the impact on waiting times, a primary objective of the surgical hub initiative and NHS policy more generally. Our analysis extends this evidence by including data up to September 2024, specifically assessing the impact on waiting times and focusing exclusively on hip and knee replacement procedures.

Most of the quantitative research on similar initiatives, such as the treatment centre programme, has relied on cross-sectional comparisons [8,10]. This prior research found that patients treated in NHS treatment centres had lower LOS compared to NHS hospitals. This study provides further evidence for this effect; using a quasi-experimental design, we found that length of stay was reduced in trusts following the implementation of a surgical hub, although we note suggestive evidence of pre-existing declines in LOS in hub-adopting trusts, which may bias this estimate.

Other quasi-experimental studies of new surgery providers have focused on private sector providers in the UK [14] or ambulatory surgical centres in the US [22] and shown substantial volume increases (12% after the introduction of private sector providers in the UK).

### Strengths and Limitations

A primary strength of this study was its quasi-experimental design using recent advances in difference-in-differences estimation. By analysing the staggered implementation of surgical hubs across NHS trusts, the study provided evidence of their impact that controls for both time-invariant local factors and common temporal factors affecting all providers. The event study approach provided important insights into implementation effects, revealing that volume increases appeared immediately following hub opening, while length of stay reductions emerged more gradually.

The dataset covered NHS acute trusts across England over a decade, with comprehensive coverage and substantial statistical power. Our focus on standardised orthopaedic procedures enabled like-with-like comparisons across trusts.

This analysis faces several limitations. First, although it included casemix measures in a sensitivity analysis, it relied on administrative data and lacked clinical detail about case complexity, potentially missing shifts in patient selection that could have influenced outcomes. There was some suggestive evidence that trusts which opened surgical hubs already had falling LOS prior to hub opening, suggesting the estimated impact of hub opening on LOS could be affected by bias.

Our trust-level analysis did not account for regional dynamics, including potential patient diversion between neighbouring trusts or the influence of independent sector provision. Our analysis examined outcomes of hub opening within two years of opening, which may differ from longer-term impacts.

Another limitation of our study is that it did not assess patient perceptions of length of stay; while shorter stays offer efficiency benefits, some patients may not prefer a reduced length of stay in all contexts. Other important dimensions of healthcare quality remain unexplored, including patient-reported outcomes, complications, readmissions, and equity of access across socioeconomic and demographic groups. Furthermore, this study is limited by not assessing whether the drive for efficiency in surgical hubs creates a conflict with surgical training, as the model may not support the necessary time and supervision for trainees. The ongoing MEASURE study will address these research questions [25].

## Conclusions

The findings of this study suggest that the implementation of surgical hubs has been effective in increasing surgical throughput for hip and knee replacement procedures, supporting their role as a key strategy for expanding capacity within the NHS. With further investment planned, policymakers can reasonably expect additional gains in surgical activity. However, the absence of a significant reduction in waiting times in the short run indicates that expanded capacity alone may not be sufficient to address the backlog of elective procedures.

The finding that volume increased substantially without a commensurate fall in waiting time aligns with some findings from related literature in the UK and US [14, 22], suggesting that the introduction of new facilities may expand demand in local areas. Consequently, we should recognise that policies designed to reduce waiting times primarily through capacity expansion may struggle to achieve their immediate objectives.

The observed reduction in length of stay may reflect the desired improved operational efficiency from hub implementation, although our research cannot rule out other mechanisms. Overall, surgical hubs seem to be an effective intervention to increase elective surgical volume, but we cannot be sure if this is through a simple increase in capacity or productivity improvements.

Future research from the MEASURE study will provide a rigorous evaluation of whether surgical hubs deliver productivity gains beyond capacity expansion and test for spillover effects on non-hub patients [25].

## Data Availability

The authors cannot share the data underlying this study as it was provided under a data sharing agreement from NHS England. Hospital Episode Statistics (HES) data and information on surgical hub opening dates can be requested from NHS England.

## List of abbreviations

APC: Admitted Patient Care
ASC: Ambulatory surgical centre
ATT: Average treatment effect on the treated
BJS: Borusyak, Jaravel and Spiess
CI: Confidence interval
COVID-19: Coronavirus disease 2019
HES: Hospital Episode Statistics
ICD-10: International Classification of Diseases, 10th Revision
IQR: Interquartile range
LOS: Length of stay
MEASURE: Mixed Methods EvAluation of the high-volume low-complexity Surgical hUb pRogrammE
NHS: National Health Service
NIHR: National Institute for Health and Care Research
OLS: Ordinary least squares
OPCS: Office of Population Censuses and Surveys Classification of Interventions and Procedures
SD: Standard deviation
T&O: Trauma and orthopaedic
TC: Treatment centre
TWFE: Two-way fixed effects

## Declarations

### Ethics approval and consent to participate

Ethics approval was not required for this study. We received exemption from ethics approval from the Chair of the Research Governance Committee for the Department of Health Sciences at the University of York. NHS England data were accessed under a data sharing agreement and held in the University of York Data Safe Haven, an ISO 27001-certified environment for handling sensitive data. The study used pseudonymised secondary administrative data and did not involve direct recruitment of participants. Consent to participate was therefore not required. Results were reported at aggregated levels, with small numbers suppressed in accordance with NHS England guidance.

### Consent for publication

Not applicable. The manuscript does not contain identifiable individual-level information.

### Availability of data and materials

The data underlying this study cannot be shared by the authors because they were provided under a data sharing agreement with NHS England. Hospital Episode Statistics data and information on surgical hub opening dates can be requested from NHS England, subject to the relevant application and approval processes.

### Authors’ contributions

PS, JA and JW conceived and designed the study. JW performed the statistical analyses. PS and JW drafted the manuscript. ZA, AC, ASt, ASc, NG, SD, KB, AR, ACA, KGB and PL critically revised the manuscript. All authors read and approved the final manuscript and agreed to be accountable for all aspects of the work. PS and JW had full access to the study data and take responsibility for the integrity of the data and the accuracy of the analysis. PS is the guarantor.

## Acknowledgements

This work uses data provided by patients and collected by the NHS as part of their care and support. Hospital Episode Statistics are copyright © 2014–2024 NHS England and are reused with the permission of NHS England. All rights reserved.

The study was undertaken in the University of York Data Safe Haven, an ISO 27001-certified environment for handling sensitive data. We are grateful for the support of the York Data Safe Haven team and Research Computing team.

Other members of the MEASURE team not separately listed as authors include Ben Ayres, Firoza Davies, Ahmed Saad, Helen Anderson and Holly Essex. We also thank the patients and members of the public who contributed to the wider MEASURE project and to the interpretation of this study.

## Footnotes

1 1000 procedures over 10 years is 23.8 procedures per quarter, this compares to a mean of 228 procedures/quarter for trusts with hubs opening or 154 procedures/quarter for trusts without hubs opening.

