## Appendix for "Effect of NHS surgical hubs on elective hip-and-knee replacement volume, length of stay and waiting times: national longitudinal difference-in-differences study"

### Appendices

**Appendix A: Methodology Details**

*BJS model*

Treatment effects are estimated using the imputation estimator of Borusyak, Jaravel and Spiess (2024). The approach proceeds in three steps. First, using untreated observations only (trusts where no surgical hub is open during the analytical period), we fit a model of untreated potential outcomes with trust and quarter fixed effects:

$$Y_{it}=\alpha_{i}+\gamma_{t}+u_{it}for untreated (i,t).$$

This yields predictions $\hat{Y}_{it}(0)$for all observations. Second, for intervention trusts we form trust-quarter level effects

$$\hat{\tau}_{it}=Y_{it}-\hat{Y}_{it}(0).$$

Third, we average $\hat{\tau}_{it}$across **treated observations** that share the same relative event time $k=t-E_{i}$to obtain event-time effects $\hat{\tau}(k)$, where, $E_{i}$denotes the first quarter in which trust $i$’s hub is operating.

We report $\hat{\tau}(k)$ for a symmetric window of −8 to +7 quarters around hub opening. Standard errors are obtained from the influence-function formula in BJS, clustered at trust level. Estimates are produced with Stata command did_imputation, and event-study graphs with event_plot.

*Two-way fixed effects*

The model is given by:

*Y_pt_ = β_0_ + β_1_SurgicalHub_pt_ + λ_p_ + α_t_ + λ_p_* ✕ *t* *+ Є_pt_*

Where *p* indexes the NHS trust and *t* indexes time in quarters. Trusts opening the surgical hubs are expressed by a dummy variable *SurgicalHub_pt_*, which equals one if trust *p* opens the surgical hub in or after quarter *t*, and zero otherwise. The dependent variable, *Y_pt,_* represents the outcome of interest for trust *p* in time *t*. *λ_p_* is a vector of NHS trust dummies which account for systematic unobserved factors that differ across hospitals but do not vary over time; *α_t_* is a vector of quarter dummies which account for aggregate change in outcomes over time. We also include the trust-specific time trends, (*λ_p_* ✕ *t*), to account for all unobserved confounding factors which vary linearly for every specific trust. *Є_pt_* is the error term. We estimate the equation using ordinary least squares (OLS) and cluster standard errors at the trust level. Our estimate of interest is *β_1_*, which identifies the average treatment effect on the treated (ATT) of the introduction of surgical hubs on the surgical volume, length of stay and waiting times. Specifically, the identification of the effect comes from whether surgical hubs lead to deviations from pre-existing trust-specific trends.

**Appendix B: Additional Results**

**Table B1: Impacts of surgical hub opening on trust-level outcomes (two-way fixed effect models, with trust specific time trends)**

|  | Procedures | LOS | Waiting Time |
| --- | --- | --- | --- |
| Post-hub | 48.90*** | -0.32** | -16.72 |
|  | [13.99, 83.80] | [-0.60, -0.04] | [-39.59, 6.15] |
| Outcome Mean | 182 | 3.9 | 168 |
| Observations | 3,158 | 3,158 | 3,097 |

Notes: 95% confidence intervals in brackets. *p < 0.1; **p < 0.05; ***p < 0.01.

**Table B2: Impacts of surgical hub opening on trust-level outcomes (two-way fixed effect models, without trust specific time trends)**

|  | Procedures | LOS | Waiting Time |
| --- | --- | --- | --- |
| Post-hub | 30.87* | -0.32*** | -17.80 |
|  | [-4.64, 66.39] | [-0.56, -0.09] | [-40.91, 5.30] |
| Outcome Mean | 182 | 3.9 | 168 |
| Observations | 3,158 | 3,158 | 3,097 |

Notes: 95% confidence intervals in brackets. *p < 0.1; **p < 0.05; ***p < 0.01.

**Table B3: Impacts of surgical hub opening on trust-level outcomes (two-way fixed effect models, with trust specific time trends, without enforcing sample restrictions)**

|  | Procedures | LOS | Waiting Time |
| --- | --- | --- | --- |
| Post-hub | 36.43** | -0.28* | -18.98* |
|  | [3.31, 69.56] | [-0.56, 0.00] | [-40.01, 2.06] |
| Outcome Mean | 170 | 3.9 | 163 |
| Observations | 4,223 | 4,223 | 4,146 |

Notes: 95% confidence intervals in brackets. *p < 0.1; **p < 0.05; ***p < 0.01.

**Table B4: Impacts of surgical hub opening on trust-level outcomes (BJS ATT without enforcing sample restrictions)**

|  | Procedures | LOS | Waiting Time |
| --- | --- | --- | --- |
| Treatment Effect | 29.74** | -0.21** | -12.10 |
|  | [6.92, 52.57] | [-0.38, -0.04] | [-28.36, 4.16] |
| Outcome Mean | 155 | 3.9 | 170 |
| Observations | 3,264 | 3,264 | 3,120 |

Notes: 95% confidence intervals in brackets. *p < 0.1; **p < 0.05; ***p < 0.01.

**Table B5: Impacts of surgical hub opening on trust-level outcomes (TWFE with casemix controls and with trust specific time trends)**

|  | Procedures | LOS | Waiting Time |
| --- | --- | --- | --- |
| Treatment Effect | 47.74*** | -0.30** | -18.48 |
|  | [13.38, 82.10] | [-0.56, -0.04] | [-40.83, 3.87] |
| Share of Female | 5.03 | 0.05 | -13.98 |
|  | [-49.69, 59.76] | [-1.02, 1.13] | [-63.89, 35.93] |
| Mean Age | 0.45 | 0.10*** | 0.69 |
|  | [-1.56, 2.46] | [0.05, 0.13] | [-1.53, 2.91] |
| Share of Less Deprived | -34.22 | -0.00 | -89.86** |
|  | [-82.20, 13.75] | [-1.04, 1.04] | [-157.15, -22.57] |
| Share of Emergency Exp. | -81.61** | 2.31*** | -121.88*** |
|  | [-145.67, -17.56] | [1.34, 3.28] | [-181.47, -62.29] |
| Mean Charlson Comorbidity | -5.61 | 0.42*** | -0.48 |
|  | [-20.87, 9.65] | [0.11, 0.73] | [-14.88, 13.92] |
| Share of Knee Replacement Patients | 23.52 | -0.79** | 71.04*** |
|  | [-22.18, 69.22] | [-1.54, -0.03] | [19.38, 122.71] |
| Outcome Mean | 182 | 3.90 | 168 |
| Observations | 3,158 | 3,158 | 3,097 |

Notes: 95% confidence intervals in brackets. *p < 0.1; **p < 0.05; ***p < 0.01. Controls include average age, share of female patients, share of less deprived patients, mean of Charlson comorbidity index, knee replacement share, and mean of prior emergency admissions at trust-quarter level.

**Table B6: Impacts of surgical hub opening on trust-level outcomes, excluding 2020/21 (BJS ATT)**

|  | Procedures | LOS | Waiting Time |
| --- | --- | --- | --- |
| Treatment Effect | 41.79*** | -0.29*** | -25.74*** |
|  | [18.82, 64.77] | [-0.50, -0.08] | [-43.66, -7.82] |
| Outcome Mean | 172 | 3.8 | 173 |
| Observations | 2,128 | 2,128 | 2,099 |

Notes: 95% confidence intervals in brackets. *p < 0.1; **p < 0.05; ***p < 0.01.
